# Work-related Musculoskeletal Pain Among Factory Workers in Kigali: Prevalence and Coping Strategies

**DOI:** 10.64898/2026.08.22.26361118

**Authors:** Pacifique Dusabeyezu, Jean Baptiste Sagahutu, Joanitah Kemigisha

**Author notes:** Corresponding Author: Pacifique Dusabeyezu.

## Abstract

**Background:** Work-related musculoskeletal pain (MSP) is a prevalent concern among factory workers, impacting productivity, absenteeism, and overall quality of life. Factory workers, who perform repetitive tasks and engage in physically demanding work, are particularly susceptible to work-related MSP.

**Methods:** This descriptive cross-sectional quantitative study included 148 factory workers (29 females and 119 males) from two factories in Kigali, surveyed between November 2023 and January 2024. Data were collected using a structured questionnaire that included demographic items, the Modified Nordic Musculoskeletal Questionnaire (M-NMQ) to assess work-related musculoskeletal pain (MSP) symptoms in nine body regions over the previous 12 months and seven days, and questions regarding interference of symptoms with normal work activities. The Coping Strategies Questionnaire (CSQ) was administered to identify coping practices among factory workers. Data analysis comprised descriptive statistics, including mean, frequency, Wilson 95% confidence intervals, and percentage values, calculated using SPSS version 25.0.

**Results:** Participants were predominantly male (119/148, 80.4%), and 98/148 (66.2%) had worked at the factory for more than five years. Overall, 113/148 reported symptoms in at least one body region during the previous 12 months (76.4%, 95% CI 68.9-82.5). The lower back (90/148, 60.8%), shoulders (71/148, 48.0%), neck (61/148, 41.2%), and knees (58/148, 39.2%) were most frequently affected. Symptoms prevented normal work for 41/148 (27.7%, 95% CI 21.1-35.4), and 57/148 reported symptoms during the previous seven days (38.5%, 95% CI 31.1-46.5). Among 113 participants with 12-month symptoms, the most frequent Always responses were seeking medical help (78/113, 69.0%), rest and recovery (75/113, 66.4%), and medication use (59/113, 52.2%); ergonomic adjustment was least frequent (10/113, 8.8%).

**Conclusion:** Self-reported work-related MSP symptoms were common, particularly in the lower back and shoulder, while ergonomic adjustments were uncommon. Frequent medication use highlights the need for comprehensive interventions combining ergonomic training, physiotherapy, and adequate recovery; however, these findings do not establish causation or intervention effectiveness.

## INTRODUCTION

Work-related Musculoskeletal pain is a significant health issue among factory workers, with a high prevalence rate that can interfere with activities at work and can lead to a reduction in productivity, increased absenteeism, chronic occupational disability, and reduced quality of life(QOL)[1]. Musculoskeletal pain (MSP) is the pain that affects bones, muscles, ligaments, tendons, and nerves and can be either acute or chronic[2], which are caused or aggravated primarily by the performance of work and by the effects of the immediate environment in which work is carried out. The pain associated with musculoskeletal (MSK) disorders is a common medical and socioeconomic problem worldwide. According to the World Health Organization (WHO), 20–33% of the world’s population has some form of chronic musculoskeletal pain, translating to 1.75 billion people globally[3]. It is also referred to as work-related musculoskeletal disorders (WMSDs), which can cause symptoms such as pain, numbness, and tingling, as well as reduced worker productivity, lost time from work, and temporary or permanent disability[4]. They result in pain and, in some cases, disability may threaten both the future of many workers and the effectiveness of many organizations [5]. WRMSDs are the leading work-related health concern in high, middle and low-income countries, accounting for over 30% of all injuries and requiring time off work [6].

These disorders include clinical syndromes such as tendon inflammations and related conditions (tenosynovitis, epicondylitis, bursitis), nerve compression disorders (carpal tunnel syndrome, sciatica), and osteoarthritis, as well as less well standardized conditions such as myalgia, low back pain and other regional pain syndromes not attributable to known pathology. Body regions most commonly involved are the low back, neck, shoulder, forearm, and hand, although recently the lower extremity has received more attention [7]. Musculoskeletal pain is a common health issue among factory workers, with a high prevalence rate and can develop into chronic pain syndromes that can be challenging to manage worldwide [3]. Musculoskeletal disorders are among the most prevalent and the most frequently reported work-related diseases. This disorder affects all age groups and causes pain which may be mild or severe, local, or widespread [8].

Musculoskeletal disorders occur consistently throughout the working years. Several individual factors, such as age, gender, anthropometry, physical activity, strength, and psychosocial factors, including psychological job demands, decision latitude, social support, job insecurity, and work environment, are associated with the development of work-related MSP [6,8].

It appears that musculoskeletal problems are becoming more prevalent than any other type of occupational illness. Research has indicated a link between elevated rates of musculoskeletal pain and job-related hazards like frequent repetition, excessive force, and uncomfortable postures. The abundance of musculoskeletal conditions can be traced back to poor posture while standing or lifting, inadequate equipment, and the characteristics of the workplace [8].

The study done in Nigeria has found that factory workers are at a high risk of developing musculoskeletal pain due to their involvement in manual material handling, heavy lifting, repetitive movements, forceful manual exertion, and exposure to whole-body or segmental vibration[8]. Although there have been research works conducted on other occupations involved in manual material handling and physical workload, there is a lack of literature on the prevalence of work-related musculoskeletal pain among factory workers in Rwanda. This study aims to investigate the prevalence of work-related musculoskeletal pain among factory workers and the coping strategies they use to manage their pain while at work. The findings of this study will contribute to the existing body of knowledge on musculoskeletal pain and coping strategies that are used at work.

## MATERIALS AND METHODS

### STUDY SETTING

The research focused on two manufacturing companies located in Kigali city, which are Sulfo Rwanda Industries Ltd and Safintra Rwanda Ltd. Sulfo Rwanda Industries Ltd is a well-known manufacturer and supplier of fast-moving consumer goods (FMCGs) based in Kigali, Rwanda. They manufacture various personal care products, packaged drinking water, chemicals, confectionery, automotive supplies, and more. The factory workers work an 8-hour shift, and they may work extra hours in exceptional cases, which are then rewarded. Similarly, Safintra Rwanda Limited specializes in mineral-based manufacturing, specifically roofing and steel products. Their product line includes roofing materials and steel components, and their staff work for 8 hours a day. Any additional hours worked are compensated under specific conditions. These companies have physically demanding tasks that require workers to exert themselves. The study aimed to investigate the prevalence of work-related musculoskeletal pain among factory workers and identify the coping strategies they use. These settings provide unique environments for conducting such research.

### STUDY DESIGN

The research project utilized a cross-sectional descriptive quantitative study design since the findings were presented as frequency distribution tables, statistics, and percentages. This is a non-experimental design [9].

### STUDY POPULATION

The study involved both male and female factory workers recruited from selected factories in Kigali, Rwanda. Interested participants were approached by a researcher and data collection took place at a convenient time and location for the participant, such as during a lunch break or after work hours.

### SAMPLING METHOD AND SAMPLE SIZE

This study utilized a census sampling method [10] to enroll 148 factory workers, covering the entire eligible population at those two factories. This method ensured that the targeted group was comprehensively represented, making it possible to conduct a thorough investigation of work-related musculoskeletal pain prevalence and coping strategies within the selected factories.

### INCLUSION CRITERIA

This study included factory workers aged 18 years and above, with a minimum work experience of twelve months, currently employed in the selected factories within the city of Kigali.

### DATA COLLECTION PROCEDURE

After obtaining ethical approval, data collection took place on 29th and 30^th^ November 2023 at Sulfo Rwanda Industries Ltd and 24^th^ January 2024 at Safintra Rwanda Ltd. Before administering standardized questionnaires, the inclusion criteria were carefully extended to exclude individuals with a history of musculoskeletal pain before joining the factory, thereby enhancing the internal validity of the study. The researcher administered questionnaires using standardized instruments. These instruments are the Nordic Musculoskeletal Questionnaire (NMQ) and Coping Strategies Questionnaire (CSQ) to gather necessary data. Participants were given the option to complete the questionnaires during lunch or after work hours for their convenience at the factory site. Prior to data collection, informed consent was obtained from all participants. To enhance the internal validity of the study, individuals with a history of pre-existing musculoskeletal pain before joining the factory were also excluded. The Nordic Musculoskeletal Pain Questionnaire was utilized to evaluate the prevalence and characteristics of musculoskeletal pain among the participants.

### VALIDITY AND RELIABILITY

The study was conducted in China to assess the reliability and validity of the NMQ. The study aimed to evaluate the applicability of the Chinese version Nordic Musculoskeletal Questionnaire (CNMQ) in the shipbuilding industry, utilizing a sample of 550 workers from a shipyard in North China. The reliability assessment of CNMQ involved the calculation of a total Cronbach’s α coefficient of 0.86, indicating favorable internal consistency, and a split-half reliability coefficient of 0.73. The designed five dimensions demonstrated coefficients within the range of 0.48-0.84. Construct validity was evaluated through the identification of thirteen common factors, explaining 64.10% of the total variance and reflecting seven dimensions. Discriminant validity analysis revealed correlation coefficients between each dimension and total items, ranging from 0.01 to 0.83, demonstrating the questionnaire’s ability to differentiate between constructs. Despite some variations in correlation coefficients, the study concluded that the reliability and validity of the CNMQ are fair for assessing work-related musculoskeletal disorders among shipbuilding workers in China after appropriate adjustments[11].

On the other hand, another study was conducted to assess the validity and reliability of the CSQ to assess the internal consistency and reliability of a Swedish version of the CSQ among 282 individuals with long-term back pain. The Swedish CSQ demonstrated high internal consistency (alpha range: 0.7 to 0.8), comparable to the American version. Test-retest reliability, though varying (correlation range: 0.4 to 0.9), indicated the CSQ’s clinical utility in assessing work-related musculoskeletal pain coping strategies[12].

### DATA ANALYSIS

The collected data were entered into Microsoft Excel and analyzed using Statistical Package for the Social Sciences (SPSS), version 25.0. Descriptive statistics were calculated to summarize the demographic characteristics, work-related factors, prevalence of musculoskeletal pain, and coping strategies among factory workers in Kigali, Rwanda. No hypothesis tests or multivariable models were performed because the study objective was descriptive and the available variables did not measure task-level ergonomic exposures. The findings of the study were presented in the form of graphs and tables to provide a clear and concise summary of the results.

### ETHICAL CONSIDERATIONS

The study was conducted with the approval of the Institutional Review Board of the University of Rwanda’s College of Medicine and Health Sciences (Ref: CMHS/IRB/388/2023). Permission was requested from the two factories to conduct the study among their workers. Study participation was voluntary, and no financial incentives were provided to participants. All data were kept confidential and anonymous, and written consent was obtained from participants at the beginning of the study. They were informed of their right to withdraw from the study at any time without any penalty and what to expect from the study.

## RESULTS

### DEMOGRAPHIC INFORMATION OF FACTORY WORKERS

The study included 148 factory workers. The largest age group was 26–35 years, comprising 50 participants (33.8%), followed by those aged 46–56 years (41/148, 27.7%) and 36–45 years (38/148, 25.7%). Most participants were predominantly male (119/148, 80.4% and had worked at their current factory for more than five years (98/148, 66.2%) and worked a minimum of 6–8 hours per day (94/148, 63.5%). Previous musculoskeletal pain before joining the current factory was reported by 73 participants (49.3%), while 18 (12.2%) reported a pre-existing condition, injury, or trauma that could affect their musculoskeletal health (Table 1).

**Table 1:**
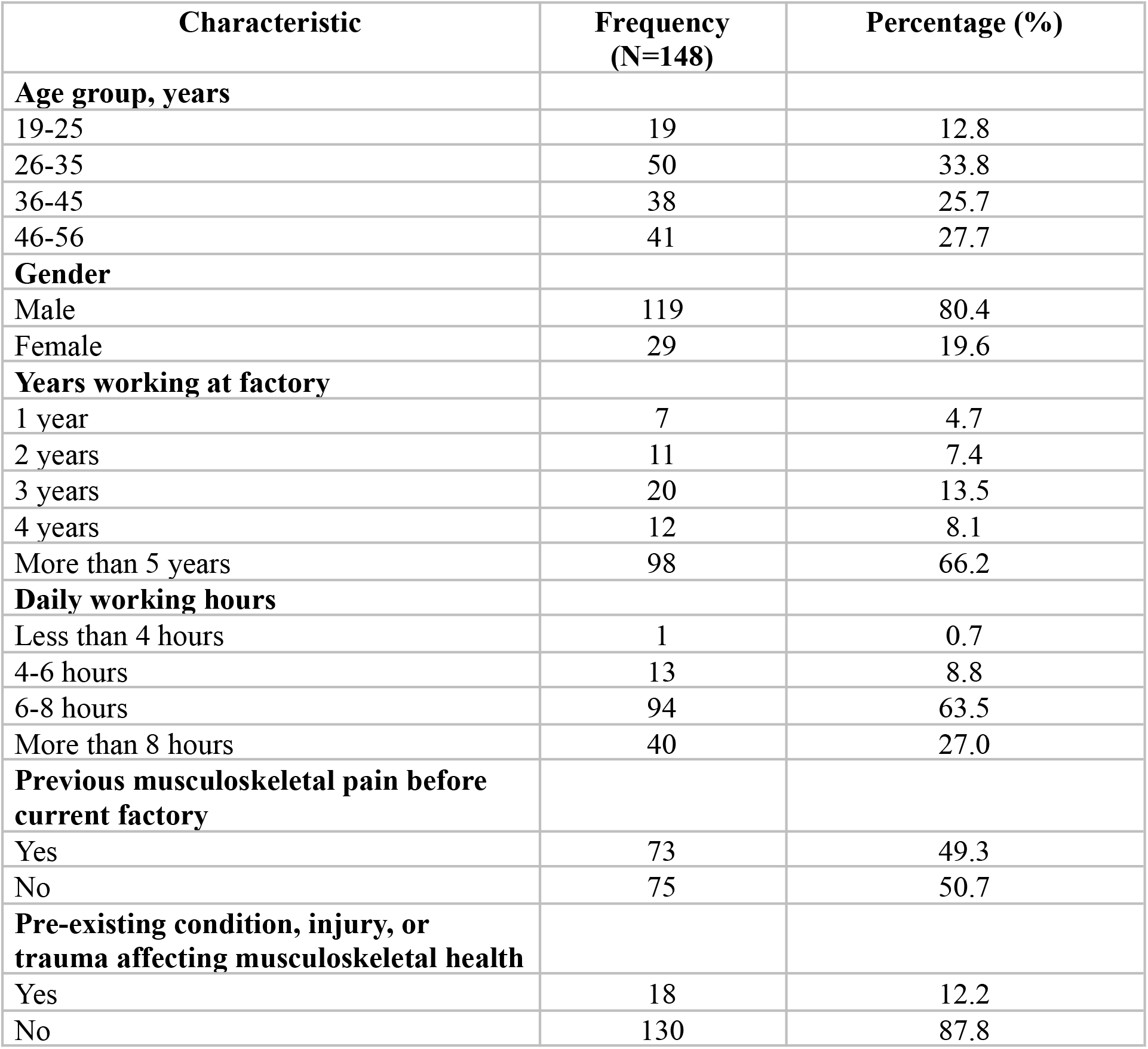
Demographic characteristics of the factory workers.

### MUSCULOSKELETAL DISCOMFORT

Overall, 113/148 participants reported work-related MSP symptoms in at least one body region during the previous 12 months (76.4%; 95% CI: 68.9–82.5). The lower back was most frequently affected (60.8%), followed by the shoulders (48.0%), neck (41.2%), and knees (39.2%). Symptoms prevented normal work in at least one region for 41/148 participants (27.7%; 95% CI: 21.1–35.4), while 57/148 reported symptoms during the previous seven days (38.5%; 95% CI: 31.1–46.5). The regional distribution of these symptoms is shown in Fig. 1.

**Fig 1.**
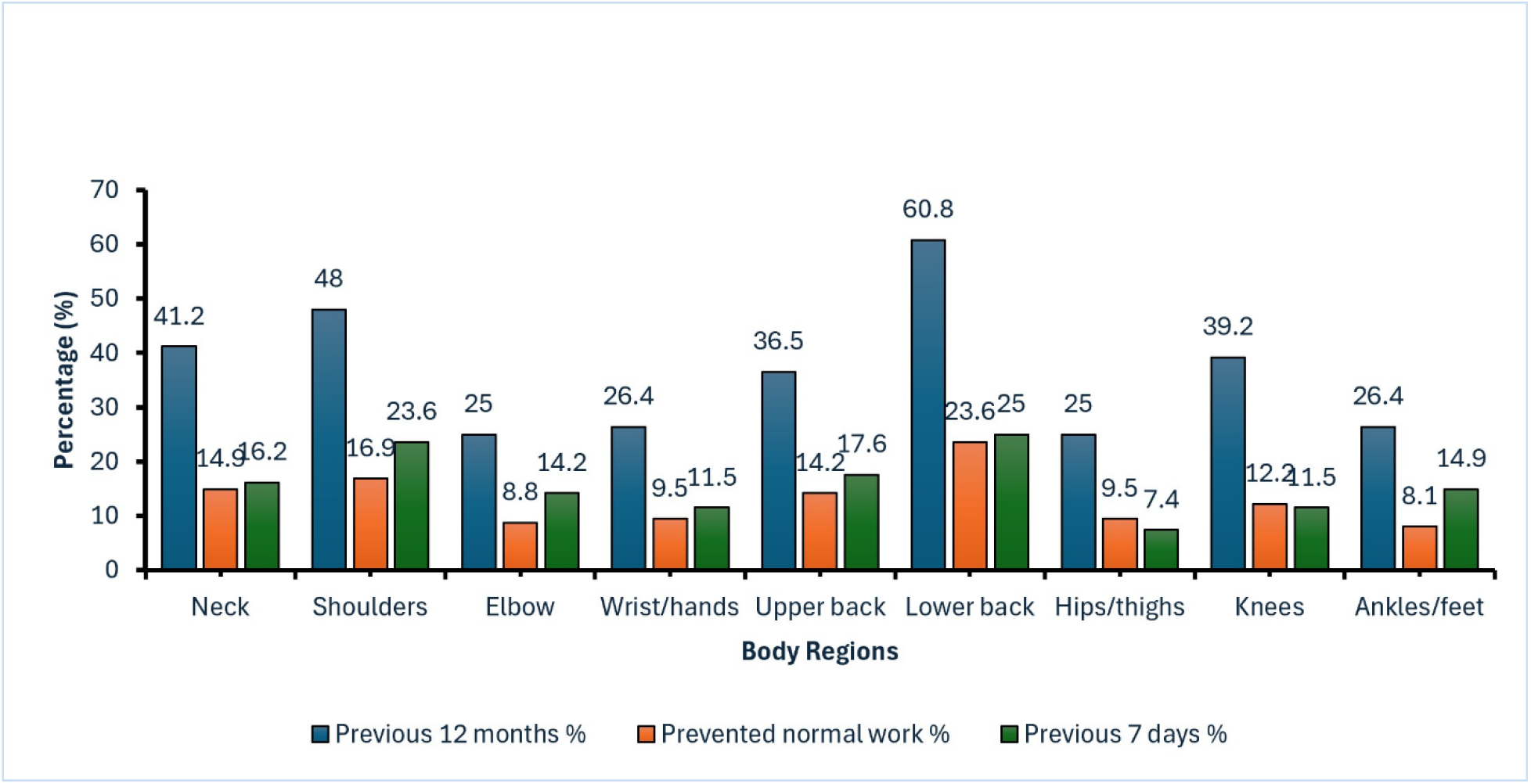
Self-reported musculoskeletal symptoms by body region among factory workers (N=148).

### COPING STRATEGIES ASSESSMENT

#### Coping Strategies employed by factory workers to manage their work-related musculoskeletal pain

Among the 113 factory workers who reported musculoskeletal symptoms in at least one body region during the previous 12 months, seeking help from a medical professional was the most frequently reported coping practice, with 78 participants (69.0%) reporting that they always sought medical help (mean score: 1.61). Prioritizing rest and recovery outside work was also commonly reported, with 75 participants (66.4%) indicating that they always used this strategy (mean score: 1.58). The use of medication was reported as always used by 59 participants (52.2%; mean score: 1.25).

Strengthening exercises or physical therapy were sometimes used by 59 participants (52.2%), whereas 26 participants (23.0%) reported always using this strategy (mean score: 0.98). Regular breaks to stretch and relax muscles were reported as always used by 22 participants (19.5%; mean score: 0.80). Ergonomic adjustment of workstations or tools was the least frequently used practice: 73 participants (64.6%) reported never using this strategy and only 10 (8.8%) reported always using it (mean score: 0.44) (Table 2)

**Table 2:**
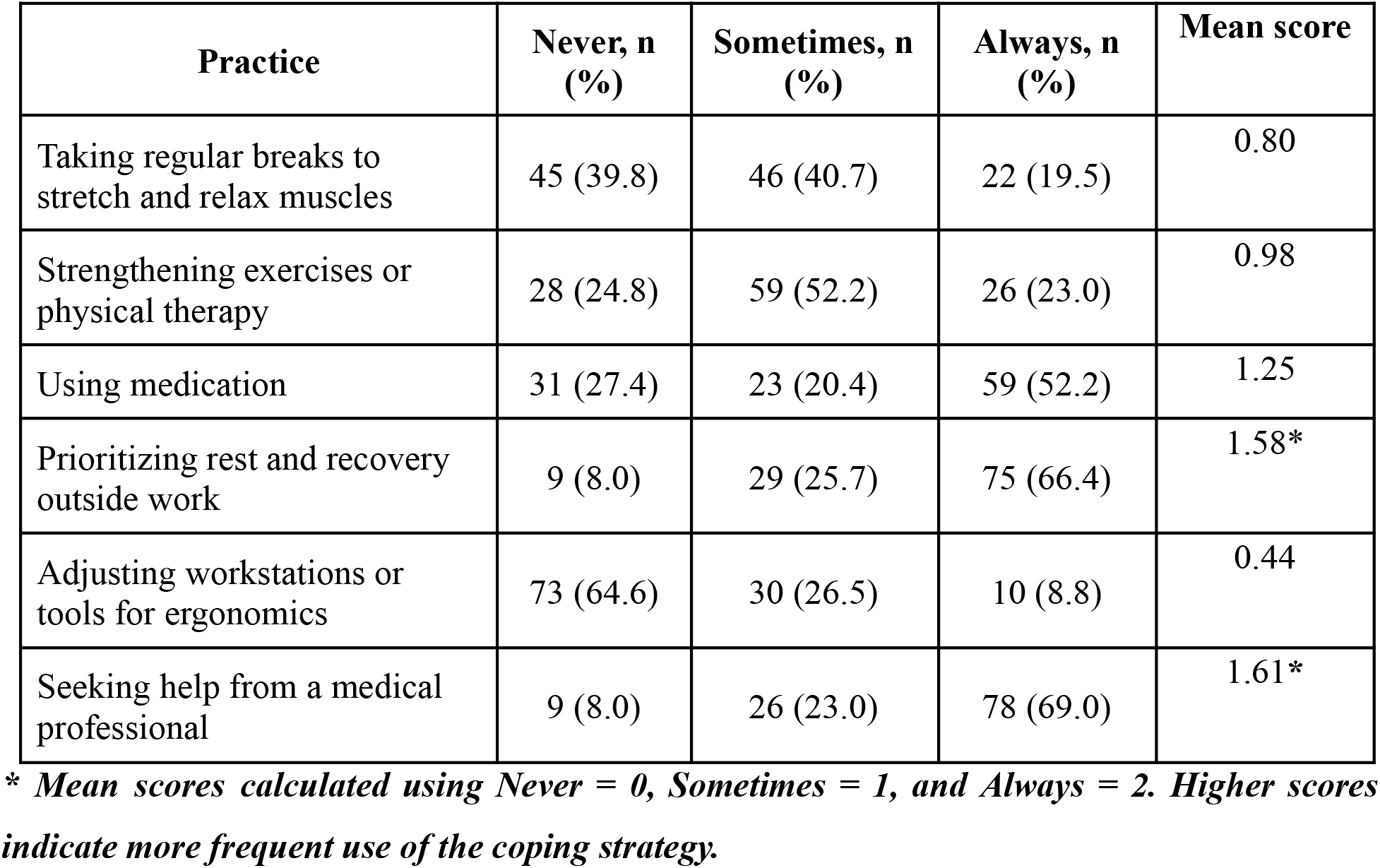
Coping Strategies employed by factory workers with at least one 12-month symptom (N = 113) to manage their work-related musculoskeletal pain symptoms.

| Practice | Never, n (%) | Sometimes, n (%) | Always, n (%) | Mean score |
| --- | --- | --- | --- | --- |
| Taking regular breaks to stretch and relax muscles | 45 (39.8) | 46 (40.7) | 22 (19.5) | 0.80 |
| Strengthening exercises or physical therapy | 28 (24.8) | 59 (52.2) | 26 (23.0) | 0.98 |
| Using medication | 31 (27.4) | 23 (20.4) | 59 (52.2) | 1.25 |
| Prioritizing rest and recovery outside work | 9 (8.0) | 29 (25.7) | 75 (66.4) | 1.58* |
| Adjusting workstations or tools for ergonomics | 73 (64.6) | 30 (26.5) | 10 (8.8) | 0.44 |
| Seeking help from a medical professional | 9 (8.0) | 26 (23.0) | 78 (69.0) | 1.61* |

*\* Mean scores calculated using Never = 0, Sometimes = 1, and Always = 2. Higher scores indicate more frequent use of the coping strategy.*

## DISCUSSION

This cross-sectional study was conducted to determine the prevalence and anatomical distribution of self-reported work-related MSP symptoms that prevented normal work, and coping practices employed by factory workers in Kigali, Rwanda. Overall, 113 of 148 participants (76.4%) reported musculoskeletal symptoms in at least one body region during the previous 12 months. Symptoms during the previous seven days were reported by 57 participants (38.5%), while 41 participants (27.7%) reported symptoms that had prevented normal work during the previous 12 months. These findings are highly consistent with Grish N et al (1), who reported a significant prevalence of musculoskeletal pain among factory workers, although this cross-sectional design does not establish whether employment at the factories caused these symptoms. The observed prevalence of work-related musculoskeletal pain (MSP) among factory workers in Kigali aligns with similar studies conducted worldwide. The study conducted in the Republic of Congo also shows the same pattern of distribution concerning the prevalence of MSDs; about 61.2% of workers had experienced MSD symptoms in at least one body region, and 25.0% were work-related musculoskeletal disorders [6].

The lower back was the most frequently affected body region, with 90 participants (60.8%) reporting symptoms during the previous 12 months. Symptoms were also frequently reported in the shoulders (71 participants, 48.0%), neck (61 participants, 41.2%), and knees (58 participants, 39.2%). This anatomical pattern is broadly consistent with studies among workers in physically demanding occupations, where lower-back, shoulder, and neck symptoms are commonly reported[13–15]. Repetitive activities, manual material handling, prolonged standing, forceful exertion, and non-neutral postures may contribute to musculoskeletal loading in factory work; however, these exposures were not measured directly in the present study and therefore cannot be evaluated as explanatory factors.

In this study, 61 participants (41. 2%)reported neck symptoms, 37 (25. 0%)reported elbow symptoms, 39 (26. 4%)reported wrist/hand symptoms, 54 (36. 5%)reported upper-back symptoms, 37 (25. 0%)reported hip/thigh symptoms, 58 (39. 2%)reported knee symptoms, and 39 (26. 4%)reported ankle/foot symptoms during the previous 12 months. During the previous seven days, neck symptoms were reported by 24 participants (16. 2%), shoulder symptoms by 35 (23. 6%), wrist/hand symptoms by 17 (11. 5%), upper-back symptoms by 26 (17. 6%), lower-back symptoms by 37 (25. 0%), hip/thigh symptoms by 11 (7. 4%), knee symptoms by 17 (11. 5%), and ankle/foot symptoms by 22 (14. 9%). However, these recall periods describe the timing of self-reported symptoms and should not be interpreted as clinical diagnoses of chronic or acute pain within their respective recall periods, rather than as measures of chronic and acute pain.

The pattern observed in this study is similar to findings from other industrial and manual-work settings, such as mining, timber, and other physically demanding worker populations where factory workers have reported a substantial burden of musculoskeletal symptoms, frequently involving the lower back, shoulders, and neck[13,14]. Although prevalence estimates vary across studies because of differences in occupations, case definitions, measurement instruments, and recall periods, the recurrent prominence of lower-back and shoulder symptoms suggests that these regions should be prioritized in workplace assessment and prevention efforts.

Among the 113 participants who reported symptoms in at least one body region during the previous 12 months, seeking help from a medical professional was the most frequently reported coping practice: 78 participants (69. 0%)reported always using this approach. Prioritizing rest and recovery outside work was also frequently reported, with 75 participants (66. 4%)reporting that they always used this strategy. Medication use was reported as always used by 59 participants (52. 2%). These results describe the self-reported practices of symptomatic workers; they do not establish whether medical help, rest, or medication was clinically appropriate or effective for managing symptoms.

Ergonomic adjustment of workstations or tools was the least frequently reported coping practice. Among participants with musculoskeletal symptoms, 73 (64.6%) reported never adjusting workstations or tools for ergonomic reasons, and only 10 (8.8%) reported always doing so. This finding may reflect limited access to adjustable equipment, ergonomic training, worker participation in workplace design, or authority to modify tasks and workstations.

Evidence from occupational-health research suggests that ergonomic strategies may reduce musculoskeletal symptoms, particularly when they are implemented as part of a broader workplace program; however, the effect of individual ergonomic interventions varies according to the work setting, the intervention components, implementation quality, and worker participation[16]. Because this study did not assess ergonomic exposures, equipment availability, or workplace practices directly, the reasons for the low use of ergonomic adjustment cannot be determined.

Regular breaks to stretch and relax muscles were always used by only 22 symptomatic participants (19.5%), whereas 45 (39.8%) reported never using this strategy. The low reported use of breaks suggests that work schedules, production demands, staffing, or workplace culture may limit opportunities for workers to pause or stretch during shifts. A Cochrane review found low- to very-low-certainty evidence that differences in work-break frequency, duration, or type may have little or no effect on musculoskeletal symptoms or disorders among healthy workers[17]. Therefore, regular breaks should not be viewed as a stand-alone solution; rather, they may be considered alongside ergonomic assessment, workload modification, worker training, and access to appropriate clinical care. Future research should assess the feasibility and effect of structured breaks in factory settings in Kigali.

Strengthening exercises or physiotherapy were always used by 26 symptomatic participants (23.0%), sometimes used by 59 (52.2%), and never used by 28 (24.8%). The relatively low proportion reporting consistent use may reflect barriers such as cost, limited availability of rehabilitation services, lack of time, low awareness of treatment options, or limited workplace support. Occupational-health literature indicates that physiotherapists can contribute to multidisciplinary programs through ergonomic assessment, worker education, therapeutic exercise, on-site management, and return-to-work support. In addition, a systematic review of workplace interventions among workers in physically demanding jobs found evidence supporting workplace strength training to reduce musculoskeletal disorders[18]. However, this study did not measure access to physiotherapy, clinical indications, exercise adherence, symptom severity, or treatment outcomes; therefore, it cannot determine whether greater use of exercise or physiotherapy would have improved outcomes in this population.

The findings should be interpreted in light of several limitations. First, the study relied on self-reported symptoms, which may have been affected by recall error and differences in how participants interpreted pain, ache, discomfort, and work interference. Second, the cross-sectional design did not establish the temporal relationship between symptoms and workplace exposures and therefore cannot demonstrate causality. Third, only two factories in Kigali were included; therefore, the results may not be generalizable to all factory workers in Kigali or Rwanda. Fourth, the study did not measure task-specific physical exposures, psychosocial working conditions, ergonomic characteristics, clinical diagnoses, or medication types. Finally, internal consistency checks identified participants who reported seven-day symptoms or symptoms preventing normal work without reporting 12-month symptoms in the same body region. These responses were retained as recorded and may have influenced the prevalence estimates.

Despite these limitations, this study provides descriptive evidence on musculoskeletal symptoms and coping practices among workers in two factories in Kigali. The lower back, shoulders, neck, and knees were commonly reported symptom sites. Among workers reporting symptoms, medical help seeking, rest and recovery, and medication use were common coping practices, whereas ergonomic adjustment, regular breaks, and consistent strengthening exercises were less frequently reported. This pattern may suggest greater emphasis on symptom management after symptoms occur than on workplace-based prevention; however, the cross-sectional data cannot establish why this pattern was observed or whether any coping practice was effective. Future studies should use longitudinal designs, including larger and more diverse workplace samples, use validated local-language instruments, and directly assess task-specific physical demands, ergonomic conditions, psychosocial work exposures, treatment access, and symptom trajectories. Such studies could clarify risk factors and evaluate the effectiveness of workplace prevention and management strategies.

## CONCLUSIONS

Self-reported musculoskeletal symptoms were common among factory workers in the two participating factories in Kigali, Rwanda, with the lower back, shoulders, neck, and knees most frequently affected. Among workers reporting symptoms, seeking medical help, prioritizing rest and recovery, and using medication were commonly reported coping practices, whereas ergonomic adjustment of workstations or tools, regular breaks, and consistent strengthening exercises or physiotherapy were less frequently reported.

These findings provide descriptive evidence on musculoskeletal symptoms and coping practices in an understudied group of factory workers in Kigali. They support the need for workplace-specific ergonomic assessment and for prospective research that evaluates task-related physical demands, ergonomic conditions, psychosocial work factors, treatment access, and the effectiveness of prevention and management strategies. Because this was a cross-sectional study based on self-reported data from two factories, the findings should not be interpreted as demonstrating that factory work caused the reported symptoms or that any coping practice was effective.

## DATA AVAILABILITY

All relevant data are within the manuscript and the analytical dataset used for this research is in the Supporting Information section.

## SUPPORTING INFORMATION

1. [Analytical dataset for musculoskeletal symptoms and coping practices among factory workers in Kigali, Rwanda](https://docs.google.com/spreadsheets/d/1Y6eK04J9L-p5qL0VzHQ0BBG3mGEBQCSU/edit?usp=sharing&ouid=110270249651574860096&rtpof=true&sd=true)

## ACKNOWLEDGMENTS

The authors thank the factory workers who participated in this study and the management and workplace personnel at Sulfo Rwanda Industries Ltd and Safintra Rwanda Ltd for facilitating data collection.

## AUTHOR CONTRIBUTIONS

Pacifique Dusabeyezu and Jean Baptiste Sagahutu contributed to the conceptualization of the study. Pacifique Dusabeyezu, Jean Baptiste Sagahutu, and Joanitah Kemigisha contributed to the methodology. Pacifique Dusabeyezu conducted formal analysis, investigation, data curation, project administration, and preparation of the original manuscript draft. Pacifique Dusabeyezu, Jean Baptiste Sagahutu, and Joanitah Kemigisha contributed to manuscript review and editing. Jean Baptiste Sagahutu and Joanitah Kemigisha provided supervision.

## REFERENCE

1. Girish N, Ramachandra K, Maiya AG, Asha K. Prevalence of musculoskeletal disorders among cashew factory workers. Arch Environ Occup Health. 2012;67(1):37–42.

2. Alghadir A, Anwer S. Prevalence of musculoskeletal pain in construction workers in Saudi Arabia. Sci World J. 2015;2015. doi:10.1155/2015/529873.

3. El-Tallawy SN, Nalamasu R, Salem GI, LeQuang JAK, Pergolizzi J V., Christo PJ. Management of Musculoskeletal Pain: An Update with Emphasis on Chronic Musculoskeletal Pain. Pain Ther. 2021;10(1):181–209.

4. Lei L, Dempsey PG, Xu JG, Ge LN, Liang YX. Risk factors for the prevalence of musculoskeletal disorders among chinese foundry workers. Int J Ind Ergon. 2005;35(3):197–204.

5. Kier KL, Ph D, Sc M. Biostatistical Applications in Epidemiology. 2011.

6. Okello A, Wafula ST, Sekimpi DK, Mugambe RK. Prevalence and predictors of work-related musculoskeletal disorders among workers of a gold mine in south Kivu, Democratic Republic of Congo. BMC Musculoskelet Disord. 2020;21(1):1–10.

7. Jahan N, Das M, Mondal R, Paul S, Saha T, Akhtar R, et al. Prevalence of Musculoskeletal Disorders among the Bangladeshi Garments Workers. SMU Med J. 2015;(2):102–113.

8. Egwuonwu U. O A, Ezeukwu AO, Ugwuoke Jeneviv A V. Prevalence of Work-Related Musculoskeletal Pain among Timber Workers in Enugu Metropolis; Nigeria. Cont J Trop Med. 2010;5(2):11–18.

9. Kesmodel US. Cross-sectional studies – what are they good for? Acta Obstet Gynecol Scand. 2018;97(4):388–393.

10. Lammer & Badia. Unit 16 : Census and Sampling Summary. UcaEdu. 2016;1–11.

11. Reliability and validity of Chinese version Musculoskeletal Questionnaire in shipbuilding industry.pdf. .

12. Jensen IB, Linton SJ. Scandinavian Journal of Coping strategies questionnaire (CSQ): Reliability of the swedish version of the CSQ. (April 2015):37–41.

13. Choobineh A, Tabatabaei SH, Mokhtarzadeh A, Salehi M. Musculoskeletal Problems among Workers of an Iranian Rubber Factory. J Occup Health. 2007;49(5):418–423.

14. Karahan A, Kav S, Abbasoglu A, Dogan N. Low back pain: prevalence and associated risk factors among hospital staff. J Adv Nurs. 2009;65(3):516–524.

15. Torgbenu EL, Nakua EK, Kyei H, Badu E, Opoku MP. Causes, trends and severity of musculoskeletal injuries in Ghana. BMC Musculoskelet Disord. 2017;18(1):349.

16. Abi Varghese AV V Panicker V. Effect of MSDs and scope of ergonomic interventions among rubber processing workers: a systematic review. Med Lav. 2022;113(4):e2022032.

17. Luger T, Maher CG, Rieger MA, Steinhilber B. Work-break schedules for preventing musculoskeletal symptoms and disorders in healthy workers. Cochrane Database Syst Rev. 2019;2019(7). doi:10.1002/14651858.CD012886.pub2.

18. Sundstrup E, Seeberg KGV, Bengtsen E, Andersen LL. A Systematic Review of Workplace Interventions to Rehabilitate Musculoskeletal Disorders Among Employees with Physical Demanding Work. J Occup Rehabil. 2020;30(4):588–612.

